# Plasma Plasmin Generation and its Determinants in Third Trimester of Pregnancy

**DOI:** 10.1101/2023.03.14.23287241

**Authors:** Helen Chioma Okoye, Theresa Ukamaka Nwagha, Joseph Tochukwu Enebe, Chilota Efobi, Oluomachi Charity Nnachi, Chikaodili J Okwor

**Affiliations:** Coagulation Unit, Department of Haematology and Immunology, College of Medicine. University of Nigeria Ituku-Ozalla campus, Enugu; Department of Obstetrics and Gynaecology, Enugu State University of Science and Technology, College of Medicine/Teaching Hospital, Parklane Enugu; Department of Haematology and Blood Transfusion, College of Health Sciences, Nnamdi Azikiwe University Nnewi campus Nnewi; Department of Haematology and Blood Transfusion, Alex Ekwueme Federal University Teaching Hospital, Abakaliki; Department of Chemical Pathology, College of Medicine. University of Nigeria Ituku-Ozalla campus, Enugu

## Abstract

**Background:** Pregnancy, especially in the third trimester, is associated with changes in the fibrinolytic system that supports clot formation and reduces hemorrhagic risk. Imbalance in the system may occur and can be assessed by plasmin generation (PG). Certain factors are known to affect PG.

**Aim:** To assess PG and its determinants during pregnancy using plasma D-dimer and plasmin-antiplasmin (PAP) complex levels.

**Methods:** Healthy pregnant women in the third trimester of pregnancy were systematically recruited. Using the ELISA method, venous blood samples were taken to assess D-Dimer and PAP complex plasma levels. IBM Statistical Package for Social Sciences (SPSS) version 21 was used for statistical analysis.

**Results:** We studied a total of 41 subjects with a mean age ±SD and gestational age ±SD of 30.68±4.69years and 34.78±3.34weeks, respectively. The mean ± SD values of the D-Dimer and PAP complex were 194 ± 24 ng/mL and 175 ± 11 ng/mL, respectively. D-Dimer and PAP complex positively correlated with age, GA, and BMI classification. However, only age was statistically significant (p-value 0.032 and 0.016, respectively). In addition, the multiple linear regression model showed that with every unit increase in age, D-Dimer and PAP complex increased by 2.0 (95% CI 0.4 – 3.6) ng/mL and 0.8 (95% CI 0.1 – 1.5) ng/mL respectively, after controlling for GA and BMI.

**Conclusion:** D-Dimer and PAP levels increased with increasing age during the third trimester of pregnancy, showing that the woman’s age is an independent determinant of PG in the third trimester.

## Introduction

Normal pregnancy is a hypercoagulable state[1,2] with a balance between coagulation and fibrinolysis, promoting pro-coagulation and limiting fibrinolysis. Pregnancy encourages thrombosis with a reduction of hemorrhagic risk, particularly during the third trimester. An increased expression and upregulation of coagulant factors, fibrinogen, and antifibrinolytic proteins [3] prepare the normal pregnant female body for the acute and extreme hemostatic challenge resulting from the separation of the placenta[4]. Rapid and effective control of the placental bed’s haemostasis improves maternal and fetal outcomes in normal pregnancy.

Thrombin generation and breakdown of fibrinogen characterize the tilt towards pro-coagulation seen in late pregnancy. Fibrin so produced secondarily encourages fibrinolysis and promotes plasmin generation[5]. The Plasmin-Alpha-2-Antiplasmin complex (PAP) is essential in blood coagulation and fibrinolysis. It controls and regulates clot dissolution into soluble fragments and is an index of recent fibrinolytic activity[6]. Its role in counterbalancing the coagulation process in the third trimester ensures effective hemostatic control during the third stage of labour. Notwithstanding that fibrinolysis is limited in pregnancy to promote coagulation, there is still evidence of active fibrinolysis as pregnancy progresses. D dimer, a fibrin degradation product, increases as pregnancy increases[7]and indicates active fibrinolysis.

The loss of fibrinolytic activity is presumed to be the loss of plasminogen activator because the fibrinolytic response is expected when added in excess in the urokinase sensitivity test. However, the capacity for localized fibrinolytic activity is not lost because fibrinolytic degradation products are slightly raised during pregnancy.

Imbalance in coagulation and fibrinolysis may occur during pregnancy, thereby increasing either thrombotic episodes or hemorrhagic risk, especially in late pregnancy. Coagulation in normal pregnancy has been extensively studied [8,9], and there is a need to understand better the fibrinolytic process and its controls in normal pregnancy. Therefore, this study investigates PAP and D dimer levels and their determinants in normal pregnant Nigerian women.

## Methodology

### Study design and centre

The study was a cross-sectional study carried out at the University of Nigeria Teaching Hospital Ituku-Ozalla, Enugu, a federal tertiary health care institution that serves as a referral centre mainly for Enugu state, with few referrals from the neighbouring States.

### Study Subjects

The study participants included normal pregnant women attending routine antenatal visits in their third trimester of pregnancy. Participants were grouped according to their gestational age (GA) in weeks as confirmed by either an early obstetrics ultrasound scan or the last menstrual cycle (for those with a typical 28-day cycle). In addition, details of their obstetrics history were documented. Their weight (in kg) and height (in m) were used to calculate the body mass index (BMI), and subjects were grouped into categories. Women with co-morbidities, twin gestation, evidence of intrauterine growth restriction, and other obstetrics complications were excluded.

### Methods

Participants were placed in three groups according to age (<20, 20 −29, 30 - 40 years); BMI (normal 18.5 – 24.9kg/m^2^, overweight 25.0 −29.9 kg/m^2^ and obese ≥ 30.0 kg/m^2^) and GA (26 – 30 weeks, 31 – 35 weeks, 36 – 40 weeks).

Four millilitres of venous blood samples were collected from each subject into an ethylene diamine tetraacetic acid (EDTA) sample bottle. Blood samples were processed within an hour of collection by centrifuging at 1500g for 10 minutes. Platelet-poor plasma was separated, labelled, and stored in aliquots in plastic tubes in a −80°C freezer until analysis. Upon analysis, stored samples were thawed at room temperature, and plasma levels of D-Dimer and PAP were determined by ELISA using kits from Elabscience®. Assay was done as per the manufacturer’s protocol.

### Sample size calculation

We employed Cochrane’s formula, using stand normal deviate of 1.96, a 12% rate of pregnant women with standard limits of D-Dimer in a Nigerian study by Udomah et al [10]., a 10% acceptable error limit and a 10% non-response rate, a sample size of 40 was obtained.

#### Ethical Considerations

The ethical approval was from the Hospital Health Research Ethics Committee. Ethics approval number NHREC/05/01/2008B-FWA00002458-1RB00002323 The objective of the study, the risk and the benefits were explained to the participants in the language they understood. Only consenting women who met the inclusion criteria participated in the study.

### Statistical Analysis

Data were entered in Microsoft Excel and then exported to IBM Statistical Package for Social Sciences (SPSS) version 21 for statistical analysis. Descriptive statistics employed means, standard deviation, median and ranges for numerical variables, while frequencies and percentages were used to describe categorical variables. The mean PAP complex and D-Dimer comparison across demographic findings employed an independent t-test for two categories and a one-way Analysis of Variance (ANOVA) for comparisons involving more than two categories. In addition, correlation and multiple linear regression analyses were employed to explore the relationship between demographic findings (age, gestational age in weeks, and BMI) and dependent variables (PAP complex and D-Dimer). Statistical significance was set at p<0.05.

## Results

We had a total of 41 subjects. Most of them (65.9%) were between 30 to 39 years, and about half (51.2%) were in their late third trimester of pregnancy, see table 1. The mean age of the study population ± SD was 30.68±4.69 years; the median age was 30; the range was 19 – 39 years. The mean gestational age among the study population, ± SD, was 34.78±3.34 weeks; the median gestational age was 36 weeks and ranged from 28 to 40 weeks. The mean BMI of the study population ± SD was 28.84 ± 4.29 Kg/m^2^ with a median BMI of 27.70 Kg/m2 and a range of 21.70 – 39.30 Kg/m^2^. Their BMIs were classified as normal, overweight, and obese. Only 7 (17.1%) study subjects had normal BMI (figure 1).

**Table 1:** Biological and Gestational Age of pregnant women in the study

| **Variables (n= 45)** | **Frequency** | **Percentage** |
| --- | --- | --- |
| **Age category** |  |  |
| < 20 years | 1 | 2.4 |
| 20 – 29 years | 13 | 31.7 |
| 30 – 39 years | 27 | 65.9 |
| **Gestational age** |  |  |
| 26 – 30 weeks | 5 | 12.2 |
| 31 – 35 weeks | 15 | 36.6 |
| 36 – 40 weeks | 21 | 51.2 |

**Figure 1:**
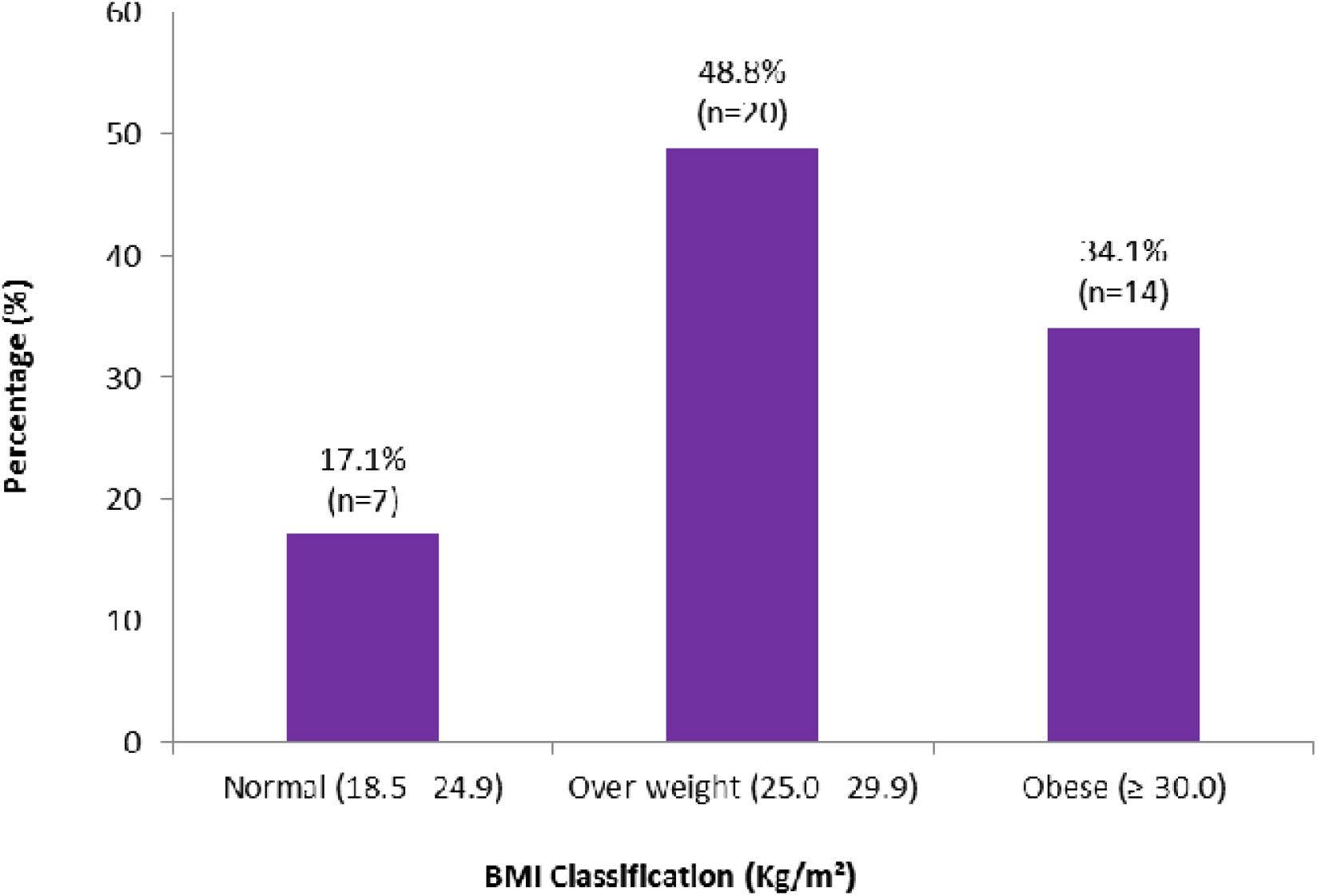
Distribution of BMI classification of study subjects.

The Mean ± SD values of D-Dimer was 194 ± 24 ng/mL, and levels ranged from 168 – 276 ng/mL. The Mean ± SD values of Plasmin-antiplasmin complex levels were 175 ± 11 ng/mL with a 156 – 200 ng/mL range among study subjects. The median values were 184 ng/mL and 172 ng/mL, respectively.

D-Dimer and Plasmin-antiplasmin complex positively correlated with age, GA, and BMI classification. However, only age was statistically significant (see table 2).

**Table 2:** Correlation between D-Dimer, Plasmin-antiplasmin complex levels and age, gestational age, BMI classification of pregnant women

| Variable | D-Dimer levels |  | Plasmin-antiplasmin complex |  |
| --- | --- | --- | --- | --- |
|  | (ng/mL) |  | (ng/mL) |  |
|  | Pearson |  | Pearson |  |
|  | Correlation<br>co-efficient (r) | p-value | Correlation<br>co-efficient (r) | p-value |
| Age in years | 0.336 | 0.032* ■ | 0.373 | 0.016* |
| Gestational age at booking (weeks) | 0.131 | 0.413 | 0.189 | 0.238 |
| BMI (Kg) | 0.070 | 0.663 | 0.137 | 0.394 |
| *Statistically significant |  | (*p*<0.05) |  |  |

The multiple linear regression model showed that with every unit increase in age, D-Dimer increases by 2.0 ng/mL (95% CI 0.4 – 3.6 ng/mL,p=0.031), see table 3. In the same way, the model showed that the PAP complex increased by 0.8 (95% CI 0.1 – 1.5ng/mL,p = 0.016) for every unit increase in age in years (see table 4). The mean D-Dimer and PAP complex levels increased across the different age group categories but did not differ significantly. In addition, there was no significant difference in D-Dimer and PAP complex levels across the different GA and BMI categories (see table 5).

**Table 3:** Multiple linear regression analysis of age, gestational age, BMI, and D-Dimer levels among pregnant women in the study

| Variables | *D-Dimer* |  |  |  |
| --- | --- | --- | --- | --- |
| | $\beta$ | 95% Confidence Interval for $\beta$ | | *p-value* |
|  |  | *Lower limit* | *Upper limit* |  |
| Age in years | 2.0 | 0.4 | 3.6 | 0.018* |
| Gestational age (weeks) | 1.1 | -1.1 | 3.3 | 0.305 |
| BMI (Kg) | 0.8 | -1.0 | 2.5 | 0.379 |

**Table 4:** Multiple linear regression analysis of age, gestational age at booking, BMI and Plasmin-antiplasmin complex levels among pregnant women in the study

| Variables | *Plasmin-Antiplasmin (PAP complex)* |  |  |  |
| --- | --- | --- | --- | --- |
| | $\beta$ | 95% Confidence Interval for $\beta$ | | *p-value* |
|  |  | *Lower limit* | *Upper limit* |  |
| Age in years | 0.8 | 0.1 | 1.5 | 0.031* |
| Gestational age at booking<br>(weeks) | -0.5 | -1.4 | 0.5 | 0.337 |
| BMI (Kg) | -0.1 | -0.9 | 0.7 | 0.763 |

\*Statistical significance $p<0.05$

**Table 5:** Comparison of mean D-Dimer, Plasmin-antiplasmin complex levels by age, gestational age, and BMI classification among pregnant women in the study

|  | D-Dimer (ng/mL) | Plasmin-antiplasmin complex |
| --- | --- | --- |
| **Variables (N= 45)** | Mean ± SD | (ng/mL)<br>Mean ± SD |
| **Age category** |  |  |
| <20 years | 178 ± - | 164 ± - |
| 20 – 29 years | 185 ± 13 | 170 ± 06 |
| 30 – 39 years | 199 ± 27 | 177 ± 12 |
| *ANOVA (p-value)* | *1.750 (0.187)* | *2.569 (0.090)* |
| **Gestational age at booking** |  |  |
| 26 – 30 weeks | 192 ± 26 | 178 ± 13 |
| 31 – 35 weeks | 190 ± 19 | 175 ± 11 |
| 36 – 40 weeks | 197 ± 27 | 174 ± 10 |
| *ANOVA (p-value)* | *0.368 (0.694)* | *0.458(0.636)* |
| **BMI classification** |  |  |
| Normal (18.0 – 24.9Kg) | 179 ± 20 | 181 ± 12 |
| Over-weight (25.0 – 29.9Kg) | 198 ± 19 | 174 ± 9 |
| Obese (≥30.0Kg) | 195 ± 29 | 173 ± 11 |
| *ANOVA (p-value)* | *1.604 (0.214)* | *1.262(0.295)* |
| *Statistically significant ($p<0.05$) | BMI - Body Mass Index | SD – |
| Standard deviation |  |  |

## Discussion

Pregnancy physiologic changes favour coagulation and reduce the hemorrhagic risk from placental separation during childbirth. Evidence shows that active fibrinolysis and plasmin generation occur in the later stages of pregnancy [5] to achieve this tilt of hemostasis balance. This study evaluated some markers of active fibrinolysis and plasmin generation in the third trimester of normal pregnancy.

Our study showed that the D-dimer levels and the plasmin-antiplasmin complex were within the physiologic reference range. Furthermore, the mean serum levels of these markers of active fibrinolysis, D dimers and PAP complex clotting factors were similar to the results reported by Choi et al [7] and Woodhams et al. [11], which showed that fibrinolytic activity rises concomitantly with increased activation of coagulation during normal pregnancy.

The level of D-dimer in our study was low compared to d-dimer levels reported in a Polish study by Siennicka et al. [12]. This study brought to the fore the need to interpret D dimer levels using pregnant reference ranges. Although the reason for the lower levels of d-dimer reported in our study could not be derived from our study, a racial disparity could be at play. Further study should be designed to answer this question.

The dip observed in the plasma levels of D-dimer at 31–35 weeks with a rise at 36–40 weeks may not be a natural dip h seen in that period of pregnancy and couldn’t be explained from our study. As previous studies like the study by Van Wersch et al [13] who reported that all makers of fibrinolysis increase during pregnancy. In addition, this dip was not reported in the study by Siennicka et al.[12] which showed a global rise of D-dimers in the third trimester though we observed the third trimester period was not categorized into weeks like in this current study. Again, our study small sample size could have been contributory.

The level of PAP complex was similar to those reported by studies like another Polish study by Uszynski *et al*. [14] this study looked at pregnant women from the third trimester through parturition. This study showed the expected physiologic changes in fibrinolysis indicators like PAP in the third trimester to reduce the hemorrhagic risk with low levels of PAP seen in the umbilical fluid. However, our study did not measure the level of PAP in the umbilical fluid as it was outside the scope of this study.

Due to the physiological changes in the haemostatic system, the interpretation levels of these fibrinolysis markers using pregnant reference ranges may be ambiguous as they could be affected by socio-demographic and obstetric factors like age, GA, and BMI, as seen in our study. From our study, the woman’s age is an independent determinant of PG in the third trimester of pregnancy. It also showed that D dimers and plasmin-antiplasmin (PAP) complex levels increased with a unit increase in maternal age in the third trimester of pregnancy. If this finding is confirmed in more extensive studies, age should be considered when interpreting D-dimer and PAP complex values in pregnancy.

Our study finds its strength in providing a quick analysis and a snapshot view of the serum levels of D-dimer and PAP complex in the third trimester. The study’s limitation was the study’s cross-sectional design; a longitudinal experimental design could have made the study more robust, especially if women were followed through parturition or compared with a non-pregnant group.

## Data Availability

All data produced in the present study are available upon reasonable request to the authors

## Data Availability

The data that support the findings of this study are available on request from the corresponding author, TUN The data are not publicly available due to [restrictions e.g., their containing information that could compromise the privacy of research participants].

## Conflict of interest

There is no conflict of interest regarding publication of this article

## Acknowledgement

the staff of the Obstetric department of UNTH and the research assistant who helped with the data collection

## Authors Contribution

HCO and TUN, contributed to the conceptualization of the study. The study was designed by HCO with input from TUN, JTE CCE, OCN, and CJO HCO, OCN, TUN, JTE, CJO, and CCE contributed to the data screening. All authors revised the manuscript for intellectual content and approved the final version.

## Notes

### Competing Interest Statement

The authors have declared no competing interest.

### Funding Statement

This study did not receive any funding

### Author Declarations

The Ethics committee/IRB of the University of Nigeria Teaching Hospital gave ethical approval for this work

